# Factors associated with the risk of upper respiratory tract bacterial infections among HIV-positive patients

**DOI:** 10.1101/2020.09.07.20189993

**Authors:** Agata Skrzat-Klapaczyńska, Marcin Paciorek, Ewa Firląg-Burkacka, Andrzej Horban, Justyna D. Kowalska

## Abstract

**Background:** The risk and characteristics of upper respiratory tract (URT) bacterial infections (URT-BI) among HIV (+) patients is understudied. We analyzed factors associated with its occurrence and the spectrum of pathogens among patients routinely followed at the HIV Out-Patient Clinic in Warsaw.

**Methods:** All symptomatic HIV (+) patients with available URT swab culture were included into analyses. Patients were followed from the day of registration in the clinic until first positive URT swab culture or last clinical visit. Cox proportional hazard models were used to identify factors associated with positive URT swabs culture (those with p<0.1 in univariate included into multivariable).

**Results:** In total 474 patients were included into the analyses, 166 with positive URT swab. In general 416 (87.8%) patients were male, 342 (72.1%) were infected through MSM contact, 253 (53.4%) were on antiretroviral therapy. Median follow-up time was 3.4 (1.3-5.7) years, age 35.2 (30.6-42.6) years and CD4+ count 528 (400–685) cells/µl. The most common pathogens were *S. aureus* (40.4%) and *S. pyogenes* (13.9%) (Table 1). Patients with URT-BI were more likely to be MSM (68.5% vs 78.9%; p<0.016), have detectable viral load (20.9% vs 12.0%; p<0.0001) and CD4+ cell count <500 cells/µl(55.2% vs 39.0%; p=0.003) (Table 2). In multivariate survival analyses detectable viral load (HR3.13; 95%Cl: 2.34-4.19) and MSM (1.63;1.09-2.42) were increasing, but older age (0.63;0.58-0.69, per 5 years older) and higher CD4+ count (0.90;0.85-0.95, per 100 cells/µl) decreasing the risk of URT-BI (Table 2).

**Conclusions:** URT BI are common among HIV (+) positive patients with high CD4+ count. Similarly to general population most common patogens are *S. aureus* and *S. pyogenes*. Risk factors identified in multivariate survival analysis indicate that younger MSM patients with detectable HIV viral load are at highest risk. In clinical practice this group of patients requires special attention.

## Introduction

The use of combination antiretroviral therapy (cART) improved the prognosis of HIV-infected patients significantly, minimizing the incidence of opportunistic diseases and significantly improving survival (1-3). Although current cART schemes show high efficacy in inhibiting viral replication, HIV elimination is still not possible (4). The use of cART allows partial reconstruction of the immune system, however the minimum HIV replication, even during effective therapy, is responsible for maintaining the activation of the immune system (5). This condition is associated with an increased risk of non-AIDS-defining diseases, such as malignant tumors, cardiovascular diseases or infections (6-8). It was observed that despite the use of cART, the frequency of deaths caused by non-AIDS-definding bacterial infections in HIV-positive population has not changed, moreover the analysis from the EuroSIDA study showed that mortality caused by non-AIDS-definding infections does not decrease with time on cART (9).

The risk and characteristics of upper respiratory tract (URT) bacterial infections (URT-BI) among HIV-positive patients is understudied. Here we analyzed factors associated with URT-BI occurrence and the spectrum of pathogens among patients routinely followed at the HIV Out-Patient Clinic in Warsaw.

## Material and methods

Electronic database of the HIV Out-Patient Clinic in Warsaw collects all medical information on patients routinely followed since 1994. The study included patients who reported to the clinic after January 1, 2007, due to the prospective management of the electronic database from that date. All symptomatic HIV-positive patients with available URT swab culture were included into analyses. Patients were followed from the day of registration in the clinic until first positive URT swab culture or last clinical visit.

For the detection of pathogens from the upper respiratory tract the routine following culture supplements were used: lamb blood agar and Schaedler’s medium as well as isolation culture media: Chapman’s substrate and MacConkey substrate. For HIV RNA identification Abbott RealTime HIV-1 Test was used. For the determination of CD4/CD8, blood samples were collected by venipuncture directly into a sterile BD Vacutainer® EDTA (ethylenediaminetetraacetic acid) tube. Identification, determination of percentages and absolute numbers of mature human T (CD3 +), helper / inducer (CD3 + CD4 +) T lymphocytes and suppressor / cytotoxic (CD3 + CD8 +) T lymphocytes in whole blood were performed using a single test tubes - BD Tritest - tri-color direct immunofluorescent reagent. Antibodies were stained with: CD4 fluorescein chlorocellulose (FITC) / CD8 phycoerythrin (PE) / CD3 peridine (PerCP).

In statistical analyses non-parametric tests were used for group comparison as appropriate. Kaplan-Meyer curves were used to investigate the time to first URT-BI in those who reported to clinic after January 1, 2007. The follow-uptime was calculated from the day of registration in the clinic until first positive URT swab culture or last clinical visit.

Univariate and multivariate Cox-proportional hazard models were used to identify factors which were associated with higher chance URT-BI. Factors considered for inclusion were age, gender, last CD4+, last CD8+, HIV RNA, antiretroviral treatment, route of infection. All analyses were performed using Statistical Analysis Software Version 9.3 (Statistical Analysis Software).

### Ethical approvals

The study was approved by the Bioethical Committee of the Medical University of Warsaw (Nr AUBE/102/15).

## Results

In total 474 patients were included into the analyses.

In general 416 (87.8%) patients were males and 58 (12,2%) patients were females. In total 342 (72.1%) patients were infected through MSM contact, 88 (18,6%) through heterosexual contact. Among the studied population 253(53.4%) patients were on antiretroviral therapy, 338 (71,3%) patients had undetectable viral load. Median follow-up time was 3.4 (1.3-5.7) years, age 35.2 (30.6-42.6) years and CD4+ count 528 (400-685) cells/µl (Table 2).

One hundred and sixty six patients had positive URT swab culture. The most common pathogens were *S. aureus* (40.4%) and *S. pyogenes* (13.9%) (Table 1).

**Table 1.** Pathogens identified in upper respiratory tract swab cultures.

| Identified pathogen | N | % |
| --- | --- | --- |
| Total number | 166 | 100 |
| 2 types of bacteria | 10 | 6 |
| 3 types of bacteria | 1 | 0,6 |
| Citrobacter spp | 1 | 0,6 |
| Enterobacter aerogenes | 1 | 0,6 |
| Enterobacter cloace | 2 | 1,2 |
| Enterococcus agglomerens | 1 | 0,6 |
| Escherichia coli | 2 | 1,2 |
| Haemophilus influenzae | 11 | 6,6 |
| Haemophilus parainfluenzae | 5 | 3,6 |
| Klebsiella pneumoniae | 10 | 6 |
| Moraxella catarrhalis | 1 | 0,6 |
| Proteus mirabilis | 1 | 0,6 |
| Serratia marcescens | 1 | 0,6 |
| Streptococcus group C, F, G | 15 | 9,02 |
| Staphylococcus aureus | 67 | 40,4 |
| Staphylococcus epidermidis | 1 | 0,6 |
| Staphylococcus haemolyticus | 1 | 0,6 |
| Streptococcus agalactiae | 1 | 0,6 |
| Streptococcus pneumoniae | 9 | 5,4 |
| Streptococcus pyogenes | 23 | 13,9 |
| Streptococcus viridans | 1 | 0,6 |

**Table 2.** Baseline characteristics for the groups of patients with positive and negative upper respiratory tract culture

| Characteristic<br>Total N=474 | N total | Positive upper<br>respiratory tract<br>culture<br>N=166 | Control group<br>N=308 | P value |
| --- | --- | --- | --- | --- |
|  | N(%) |  |  |  |
| Female | 58 (12,2) | 13 (7,8) | 45 (14,6) | 0.03 |
| Male | 416 (87,8) | 153 (92,2) | 263 (85,4) |  |
| Risk group |  |  |  | 0.01 |
| Heterosexual | 88(18,6)) | 18 (10,8) | 70 (22,7) |  |
| MSM | 342(72,1) | 131 (78,9) | 211(68,5) |  |
| IDU | 33 (6,9) | 12 (7,2) | 21(6,8) |  |
| Unknown | 11 (2,3) | 5 (3) | 6 (1,9) |  |
| ARV | 221 (46,6) | 78(47) | 143 (46,4) |  |
| No ARV | 253 (53,4) | 88(53) | 165(53,6) | 0.90 |
| Detectable viral load | 136 (28,7) | 99 (20,9) | 37 (12) | <0.0001 |
| Undetectable viral load | 338 (71,3) | 67 (40,3) | 271 (88) |  |
| Baseline CD4 |  |  |  | 0.003 |
| <350 cells/ml | 75 (15,9) | 31 (18,8) | 44(14,3) |  |
| 350-500 cells/ml | 136 (28,7) | 60 (36,4) | 76 (24,7) |  |
| >500 cells/ml | 262 (55,4) | 74 (44,8) | 188 (61) |  |
|  | Median (IQR) |  |  |  |
| Age at registration in years | 35.2 (30.6-41.6) | 31.6 (26.7-36.9) | 37.1 (32.5-44.5) |  |
| In care from 2007 | 3.43 (1.34-5.74) | 0.70 (0.21-1.84) | 4.66 (3.30-7.00) |  |
| Baseline CD4 | 528 (400-685) | 483 (374-604) | 559 (435-716) |  |
| Baseline CD8 | 851 (628-1122) | 941(674-1274) | 809(600-1051) |  |

Patients with URT-BI were more likely to be MSM (68.5% vs 78.9%; p< 0.016), have detectable viral load (20.9% vs 12.0%; p< 0.0001) and CD4+ cell count < 500 cells/µl (55.2% vs 39.0%; p = 0.003) (Table 2).

Probability of a positive URT culture according to Kaplan Meier with stratification relative to the route of infection (Log-Rank p = 0.0137) was higher in patients infected through homosexual contact and among patients, who have never admitted what was the route of HIV infection (Figure 1).

**Figure 1.**
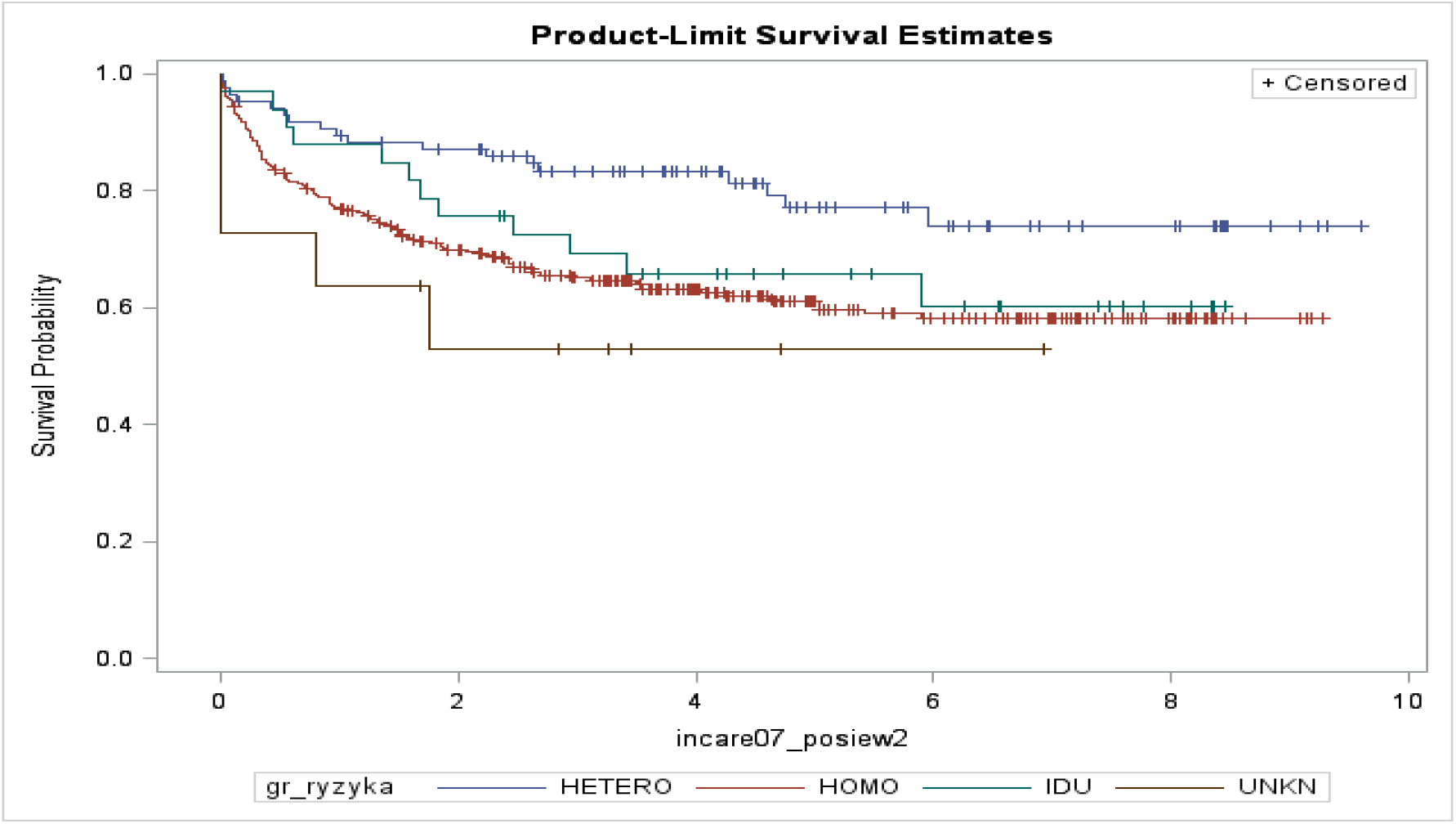
Probability of a positive culture result according to Kaplan Meier with stratification relative to the route of infection (Log-Rank p = 0.0137)

In univarinate regression models factors identified as significant were age (HR 0.90; 95% Cl:0.88-0.92), last CD4+ (0.99; 0.98-0.99), last CD8+ (1.00; 1.00-1.00) and detectable viral load (6.36; 4.64-8.73). In multivariate survival analyses detectable viral load (HR 3.13; 95% Cl:2.34-4.19) and MSM (1.63;1.09-2.42) were increasing the risk of URT-BI; whereas older age(0.63;0.58-0.69, per 5 years older) and higher CD4+ count (0.90;0.85-0.95, per 100 cells/µl) were decreasing the risk of URT-BI (Table 3).

**Table 3.** Univariate and multivariate logistic regression models for positive upper respiratory tract swab culture

|  |  | Univariate |  | Multivariate* |  |
| --- | --- | --- | --- | --- | --- |
| Variable |  | HR (95% CI) | P value | HR (95% CI) | P value |
| Age in years (per 5 years) |  | 0.90 (0.88-0.92) | <0.0001 | 0.91 (0.89-0.93) | <0.0001 |
| Last CD4+ cells/ul (per 100) |  | 0.99 (0.98-0.99) | <0.0001 | 0.99 (0.99-1.00) | 0.0005 |
| Last CD8+ cells/ul (per 100) |  | 1.00 (1.00-1.00) | <0.0001 | 1.00 (1.00-1.00) | 0.12 |
| HIV RNA >50 copies/ml |  | 6.36 (4.64-8.73) | <0.0001 | 3.13 (2.34-4.19) | <0.0001 |
| Route of inf. heterosexual | MSM | 2.10 (1.28-3.45) | 0.003 | 1.63 (1.09-2.42) | 0.01 |
|  | IDU | 1.76 (0.85-3.66) | 0.12 | 0.90 (0.57-1.43) | 0.66 |
|  | Unknown | 3.23 (1.20- | 0.02 | 2.27 (0.99- | 0.005 |
|  |  | 8.72) |  | 5.20) |  |
| Male gender |  | 1.83 (1.03-3.22) | 0.03 | - | - |
| On antiretroviral treatment |  | 0.90 (0.66-1.22) | 0,50 | - | - |
| * Adjusted for all variables significant ($p < 0.1$) in univariable analyses | | | | | |

## Discussion

In our study one in two observed patients had positive URT culture and the most common pathogen was *S. aureus* (40,4%), followed by *S. pyogenes* (13.9%).

In the general population, bacteria responsible for acute pharyngitis and tonsillitis in 5-10% is *Streptococcus pyogenes* in adults, while *group C* and *G streptococci* are found much less frequently, in 5-11% of cases (10, 11). Bacterial acute rhinosinusitis in the general population is caused in most cases by *S. pneumoniae* (26-35%) and *H. influenzae* (21-40%). The remaining microbes are responsible for less than 20% of bacterial infections and include *M. catarrhalis, S. aureus* and *S. pyogenes* (12, 13).

On the other hand in the study Verghese et al. the most common pathogens of bacterial bronchitis in HIV-positive patients were *Haemophilus influenzae, Streptococcus pneumoniae* and *Pseudomonas aeruginosa* (14). In our findings *Haemophilus influenzae* and *Streptococcus pneumoniae* were quite common – 6,6% and 5,5% respectively but *Pseudomonas aeruginosa* didn’t occur at all.

*Staphylococcus aureus* is isolated from healthy people from the anterior throat, digestive tract, urogenital tract and skin. About 30% of healthy people carry *S. aureus* in the nasopharynx, and the transmissible carrier found in about 50-60% of the general population. Increased carriage was found in patients with type 1 diabetes, hemodialyzed patients, patients with peritoneal dialysis, intravenous drug users, and HIV-infected patients (15, 16).

In 2009, a study by Shet et al. was published, which compared the colonization and subsequent infection with the etiology of *MRSA* among 107 people infected with HIV compared to the group of 52 people without HIV infection. The study was conducted for a year (2005-2006). Both groups of patients were similar. One hundred and fifty nine men surveyed were homosexuals with a mean age of 37.5. None of the respondents was hospitalized during the last year during the study. In HIV-infected patients, mean CD4 cell counts were 599 cells / μl and 74 (69.2%) of 107 infected persons regularly received antiretroviral therapy. Initially, 65 (60.7%) patients with human immunodeficiency virus were with undetectable viral load. Thus, the population of HIV-infected men was similar in terms of immunocompetence to the population of healthy males. Analysis of the total colonization frequency by *S. aureus* showed that 58.9% (63 out of 107 [95% CI, 49.4% - 67.7%]) HIV infected and 34.6% (18 out of 52 [95%] CI, 23.1% −18.2%]) of non-HIV infected subjects were colonized at least once during the study period (pp 0.004; OR: 2.7 [95% CI, 1.29-5.72]). During the study, ten people infected with HIV developed a skin and soft tissue infection with the etiology of *MRSA*, and none of the non-HIV positive was described during the same period. All persons who had an *MRSA* infection were also carriers of *MRSA* (16). In the conducted research, it was proved that colonization of the skin and mucous membranes with methicillin-resistant *Staphylococcus aureus (MRSA)* is a predisposing factor for a full-blown infection with this pathogen (17). This trend was also observed among HIV infected populations with an increased incidence of both colonization and infection (18, 19).

Patients infected with human immunodeficiency virus are particularly susceptible to *MRSA* infections because they have reduced immunity as well as due to demographic, behavioral and socio-economic factors as well as frequent exposures to the healthcare system (20, 21). In HIV-infected people, *MRSA* ranks high among the most common causes of bacterial infections, especially skin and soft tissue infections (18). In our results *Staphylococcus aureus* was the most common bacterial pathogen in MSM population, which is very interesting and requires further research.

In our study in multivariate survival analyses detectable viral load was increasing the risk of URT-BI by more than 3 times.

Reekie et al. in 2011 showed that HIV infected individuals with CD4 + cell counts greater than 350 cells / μ l and those with uncontrolled viral replication had an increased incidence of AIDS-related diseases and a slightly increased incidence of non-AIDS definding diseases. The relationship with AIDS was clear and consistent. However, after considering the complicating factors, there was also a relationship with the occurrence of non-AIDS-related events, but no differences were observed between mean and high viral load (22).

In the study from Sogaard et al. performed in 2013 there was clearly demonstrated that discontinued antiretroviral therapy is a risk factor for bacterial infections (IRR = 2.96, 95% CI: 2.03-4.32) in HIV-infected patients (23). These data are consistent with the in our study the relationship between HIV viral load> 50 copies / ml and more than 3-fold increased risk of bacterial infections.

More work has been done for serious bacterial infections in HIV-positive patients, but we can observe trends in occurrence of these events in the combination ART era. For example the incidence of pneumonia and other opportunistic infections has decreased (24, 25). Observational studies have shown that the introduction of cART led to a decrease in the frequency of hospitalizations associated with pneumonia, but the risk of pneumonia remained still six-fold higher among people with HIV than people without HIV. Similar trends are also observed in the case of bacterial meningitis and invasive pneumococcal disease (23, 26). However, bacterial pneumonia and chronic obstructive pulmonary disease (COPD) are the most prevalent lung comorbidities in people living with HIV (27). Moreover, subsequent studies have suggested that COPD, lung cancer and pulmonary hypertension may also occur with greater frequency in populations infected with HIV (28-30). The mechanisms for the observed increases in these diseases, however, are not well understood.

In our multivariate survival analyses older age was decreasing the risk of URT-BI. This is completely the opposite situation considering the lower respiratory tract infections where the factor increasing the risk of these infections is older age (31). Our results may be related to a typical epidemic trend, because young people usually stay in groups.

There are some limitations in our study which need to be mentioned. First of all data on clinical symptoms and indications for URT swab culture were not available for these analyses. This could result in underreporting of bacterial infections of the URT. However these data represent clinical practice therefore it is not unreasonable to assume that clinical symptoms were the indications for performing a swab. Secondly, it was the fact that the role of *Staphylococcus aureus* as the pathogen of the upper respiratory tract is the subject of discussion because it is a frequent component of the flora found in the nasal tracts in health. In the shared database, it was not possible to verify disease symptoms with a positive culture result. Moreover, we didn’t know if our Staphylococcus aureus was MSSA or MRSA. Therefore our findings can only be referred to population of HIV-positive patients who had URT swab culture based diagnosis.

Research works usually focus on serious bacterial infections in HIV-infected patients. There are only few publications describing URT-BI in this population. In our opinion the analysis of factors affecting the occurrence of bacterial upper respiratory tract infections in HIV-positive patients, with particular emphasis on antiretroviral treatment, remains a research challenge. Thanks to combination ARV, we can observe the aging of this population, but it is important to investigate whether URT-BI are more frequent than in the general population. Due to the fact that the detectable viral load of HIV increases the risk of upper respiratory tract bacterial infection, we should realize that the benefits of effective antiretroviral therapy also include reduction of URT-BI events.

## Data Availability

All data is available if necessary

## Acknowledgments

The above findings were presented in par at the conference HIV Drug Therapy in Glasgow 2018 as poster presentation. The study has been funded by research grant issued by the Research Development Foundation in Hospital for Infectious Diseases.

## References

1. Mocroft A, Ledergerber B, Katlama C, Kirk O, Reiss P, d’Arminio Monforte A, et al. Decline in the AIDS and death rates in the EuroSIDA study: an observational study. Lancet. 2003;362(9377): 22–9.

2. Antiretroviral Therapy Cohort C, Mocroft A, Sterne JA, Egger M, May M, Grabar S, et al. Variable impact on mortality of AIDS-defining events diagnosed during combination antiretroviral therapy: not all AIDS-defining conditions are created equal. Clin Infect Dis. 2009; 48(8): 1138–51.

3. Martinez E, Milinkovic A, Buira E, de Lazzari E, Leon A, Larrousse M, et al. Incidence and causes of death in HIV-infected persons receiving highly active antiretroviral therapy compared with estimates for the general population of similar age and from the same geographical area. HIV Med. 2007; 8(4): 251–8.

4. Thorlund K, Horwitz MS, Fife BT, Lester R, Cameron DW. Landscape review of current HIV ‘kick and kill’ cure research - some kicking, not enough killing. BMC Infect Dis. 2017; 17(1): 595.

5. Neuhaus J, Jacobs DR, Jr., Baker JV, Calmy A, Duprez D, La Rosa A, et al. Markers of inflammation, coagulation, and renal function are elevated in adults with HIV infection. J Infect Dis. 2010; 201(12): 1788–95.

6. Strategies for Management of Anti-Retroviral Therapy I, Groups DADS. Use of nucleoside reverse transcriptase inhibitors and risk of myocardial infarction in HIV-infected patients. AIDS. 2008; 22(14): F17–24.

7. Reekie J, Kosa C, Engsig F, Monforte A, Wiercinska-Drapalo A, Domingo P, et al. Relationship between current level of immunodeficiency and non-acquired immunodeficiency syndrome-defining malignancies. Cancer. 2010; 116(22): 5306–15.

8. Baker JV, Peng G, Rapkin J, Abrams DI, Silverberg MJ, MacArthur RD, et al. CD4+ count and risk of non-AIDS diseases following initial treatment for HIV infection. AIDS. 2008; 22(7): 841–8.

9. Kowalska JD, Reekie J, Mocroft A, Reiss P, Ledergerber B, Gatell J, et al. Long-term exposure to combination antiretroviral therapy and risk of death from specific causes: no evidence for any previously unidentified increased risk due to antiretroviral therapy. AIDS. 2012; 26(3): 315–23.

10. Bourbeau PP. Role of the microbiology laboratory in diagnosis and management of pharyngitis. J Clin Microbiol. 2003; 41(8): 3467–72.

11. Lindbaek M, Hoiby EA, Lermark G, Steinsholt IM, Hjortdahl P. Clinical symptoms and signs in sore throat patients with large colony variant beta-haemolytic streptococci groups C or G versus group A. Br J Gen Pract. 2005;55(517): 615–9.

12. Payne SC, Benninger MS. Staphylococcus aureus is a major pathogen in acute bacterial rhinosinusitis: a meta-analysis. Clin Infect Dis. 2007; 45(10): e121–7.

13. Brook I. Microbiology of sinusitis. Proc Am Thorac Soc. 2011; 8(1): 90–100.

14. Verghese A, al-Samman M, Nabhan D, Naylor AD, Rivera M. Bacterial bronchitis and bronchiectasis in human immunodeficiency virus infection. Arch Intern Med. 1994; 154(18): 2086–91.

15. Fournier B, Philpott DJ. Recognition of Staphylococcus aureus by the innate immune system. Clin Microbiol Rev. 2005; 18(3): 521–40.

16. Shet A, Mathema B, Mediavilla JR, Kishii K, Mehandru S, Jeane-Pierre P, et al. Colonization and subsequent skin and soft tissue infection due to methicillin-resistant Staphylococcus aureus in a cohort of otherwise healthy adults infected with HIV type 1. J Infect Dis. 2009; 200(1): 88–93.

17. Gordon RJ, Lowy FD. Pathogenesis of methicillin-resistant Staphylococcus aureus infection. Clin Infect Dis. 2008;46 Suppl 5: S350–9.

18. Crum-Cianflone NF, Burgi AA, Hale BR. Increasing rates of community-acquired methicillin-resistant Staphylococcus aureus infections among HIV-infected persons. Int J STD AIDS. 2007; 18(8): 521–6.

19. Miller M, Cespedes C, Bhat M, Vavagiakis P, Klein RS, Lowy FD. Incidence and persistence of Staphylococcus aureus nasal colonization in a community sample of HIV-infected and -uninfected drug users. Clin Infect Dis. 2007; 45(3): 343–6.

20. Imaz A, Pujol M, Barragan P, Dominguez MA, Tiraboschi JM, Podzamczer D. Community associated methicillin-resistant Staphylococcus aureus in HIV-infected patients. AIDS Rev. 2010; 12(3): 153–63.

21. Cole J, Popovich K. Impact of community-associated methicillin resistant Staphylococcus aureus on HIV-infected patients. Curr HIV/AIDS Rep. 2013; 10(3): 244–53.

22. Reekie J, Gatell JM, Yust I, Bakowska E, Rakhmanova A, Losso M, et al. Fatal and nonfatal AIDS and non-AIDS events in HIV-1-positive individuals with high CD4 cell counts according to viral load strata. AIDS. 2011; 25(18): 2259–68.

23. Sogaard OS, Reekie J, Ristola M, Jevtovic D, Karpov I, Beniowski M, et al. Severe bacterial non-aids infections in HIV-positive persons: incidence rates and risk factors. J Infect. 2013; 66(5): 439–46.

24. Palella FJ, Jr., Delaney KM, Moorman AC, Loveless MO, Fuhrer J, Satten GA, et al. Declining morbidity and mortality among patients with advanced human immunodeficiency virus infection. HIV Outpatient Study Investigators. N Engl J Med. 1998; 338(13): 853–60.

25. Le Moing V, Rabaud C, Journot V, Duval X, Cuzin L, Cassuto JP, et al. Incidence and risk factors of bacterial pneumonia requiring hospitalization in HIV-infected patients started on a protease inhibitor-containing regimen. HIV Med. 2006; 7(4): 261–7.

26. Kohli R, Lo Y, Homel P, Flanigan TP, Gardner LI, Howard AA, et al. Bacterial pneumonia, HIV therapy, and disease progression among HIV-infected women in the HIV epidemiologic research (HER) study. Clin Infect Dis. 2006; 43(1): 90–8.

27. Crothers K, Huang L, Goulet JL, Goetz MB, Brown ST, Rodriguez-Barradas MC, et al. HIV infection and risk for incident pulmonary diseases in the combination antiretroviral therapy era. Am J Respir Crit Care Med. 2011; 183(3): 388–95.

28. Diaz PT, King MA, Pacht ER, Wewers MD, Gadek JE, Nagaraja HN, et al. Increased susceptibility to pulmonary emphysema among HIV-seropositive smokers. Ann Intern Med. 2000; 132(5): 369–72.

29. Kirk GD, Merlo C, P OD Mehta SH, Galai N, Vlahov D, et al. HIV infection is associated with an increased risk for lung cancer, independent of smoking. Clin Infect Dis. 2007; 45(1): 103–10.

30. Mehta NJ, Khan IA, Mehta RN, Sepkowitz DA. HIV-Related pulmonary hypertension: analytic review of 131 cases. Chest. 2000; 118(4): 1133–41.

31. Torres A, Peetermans WE, Viegi G, Blasi F. Risk factors for community-acquired pneumonia in adults in Europe: a literature review. Thorax. 2013; 68(11): 1057–65.

